# Are Cloth Masks a Substitute to Medical Masks in reducing transmission and contamination? A Systematic Review

**DOI:** 10.1101/2020.07.27.20154856

**Authors:** Milena Santos, Darlyane Torres, Paula C. Cardoso, Nikolaos Pandis, Carlos Flores-Mir, Rita Medeiros, David Normando

**Affiliations:** Health Sciences Institute, Federal University of Pará, Belém, Pará, Brazil.; Department of Orthodontics and Dentofacial Orthopedics, School of Dentistry, University of Bern, Bern, Switzerland; School of Dentistry, Faculty of Medicine and Dentistry, University of Alberta, Edmonton, Alberta, Canada.; Tropical Medicine Group, Federal University of Pará, Belém, Pará, Brazil

**Keywords:** Masks, Pandemics, Respiratory Protective Devices, Coronavirus

## Abstract

During the COVID-19 pandemic the use of cloth masks has increased dramatically due to the shortage of medical masks. However, the efficiency of this material is controversial. We aimed to investigate the efficiency of cloth masks in reducing transmission and contamination by droplets and aerosols for the general population and healthcare workers. Electronic databases were searched without year or language restrictions. Clinical and laboratorial studies were included. The risk of bias (RoB) was assessed using an adapted quality checklist for laboratory-based studies. ROBINS-I tool and Cochrane RoB 2.0 were used to evaluate non-randomized (n-RCT) and randomized clinical trials (RCT), respectively. The quality of the evidence was assessed through GRADE tool. From the eleven studies selected, eight were laboratory-based studies, one non-randomized and one RCT supported by laboratory data. Between the evaluated fabrics only three presented a filtration efficiency >90%. Hybrid of cotton/chiffon (95%CI 95.2 to 98.8), hybrid of cotton/silk (95%CI 92.2 to 95.8) and cotton quilt (95%CI 94.2 to 97.8). A meta-analysis was not feasible due to a high methodological heterogeneity. The overall quality of evidence ranged from very low to moderate. Despite the lower efficiency compared to medical masks, laboratorial results may underestimate the efficiency of cloth masks in real life. Cloth mask efficiency is higher when made of hybrid fabrics (cotton/chiffon, cotton/silk) and cotton quilt, mainly with multiple layers. In pandemic situations any measure that can contribute to source control at the population level can have a beneficial effect. However, cloth masks are not recommended for healthcare workers.

## INTRODUCTION

According to the World Health Organization (WHO)^1^, viral diseases continue to emerge and represent a serious issue to public health. In the past few months, the COVID-19 pandemic has been the focus in scientific journals and the media. Frequent handwashing, barrier measures such as gloves, gowns and masks and isolation of suspected cases are some of the recommended procedures to reduce transmission in respiratory diseases^2^. Knowing COVID-19 is highly contagious, some experts and countries have encouraged or even implemented mandatory facial covering in public as a form of prevention^3^.

Recent studies^4,5^ reported that viral shedding of patients with the SARS-CoV-2 was higher at the time or before symptom onset. It means that a considerable portion of infected individuals with the new coronavirus are asymptomatic or pre-symptomatic patients and can transmit the virus during routine activities like speaking, coughing, or sneezing. Therefore, it is extremely important that everyone use masks, even those who did not present any symptoms.

Surgical masks, N95 respirators and similar are effective barriers that can help preventing COVID-19. However, due to the shortage of these products at the market^6^ it only should be used by healthcare workers. For the general population, the Centers for Disease Control and Prevention^7^ recommends wearing cloth mask covering in public settings, which are a simple and low coast measure that may have a big social impact.

The main objective of use these masks in public is to decrease transmission by pre-symptomatic infected individuals who continue to move freely. This is known as source control and refers to the effectiveness of blocking droplets from an infected person, when droplets expelled are not small enough to squeeze through the weave of a cotton mask^8^. When facing an epidemic episode, authorities must decide on the best actions to reduce the social impact, and source control becomes a critical matter in the debate about whether the public should wear masks^9^.

Studies suggested that in laboratory settings cloth masks are less effective than surgical masks^10,11^, but they seemed better than no protection at all^12^. Although some studies showed limited evidence regarding the use of face masks, absence of evidence is not evidence of absence^13^. With no effective vaccine or treatment, reducing the rate of infection is a urgency^14^.

Public policy makers need urgent guidance on the use of masks by the general population as a tool in combating SARS-CoV-2, based on the best available evidence. Therefore, the aim of this systematic review was to evaluate the existing evidence on the efficiency of homemade or commercial cloth masks compared to surgical masks and N95/others respirators in reducing transmission and contamination by droplets and aerosols in the general population and among healthcare workers.

## MATERIAL AND METHOD

### Search strategy and selection criteria

The present systematic review was registered in the PROSPERO database (https://www.crd.york.ac.uk/PROSPERO, CRD42020178417) and was reported according to the Preferred Reporting Items for Systematic Review and Meta-Analysis (PRISMA) guidelines (www.prisma-statement.org)^15^.Electronic searches, without date and language restrictions, were carried out according to the PECO strategy. Eight databases were used: PubMed, Scopus, Web of science, Cochrane, VHS, OpenGrey, Google Scholar and Clinical Trials (Table 1). An additional manual search was carried out in the reference list of the included articles, as well as in some hand-picked electronic journals. Alerts were received by April 30, 2020.The eligibility criteria were defined according to the PECO research strategy for clarity in resolving the question: Can homemade or commercial cloth masks be used instead of surgical masks and N95 respirators as an alternative in reducing transmission and contamination by droplets and aerosols for general population and healthcare workers?

**Table 1.**
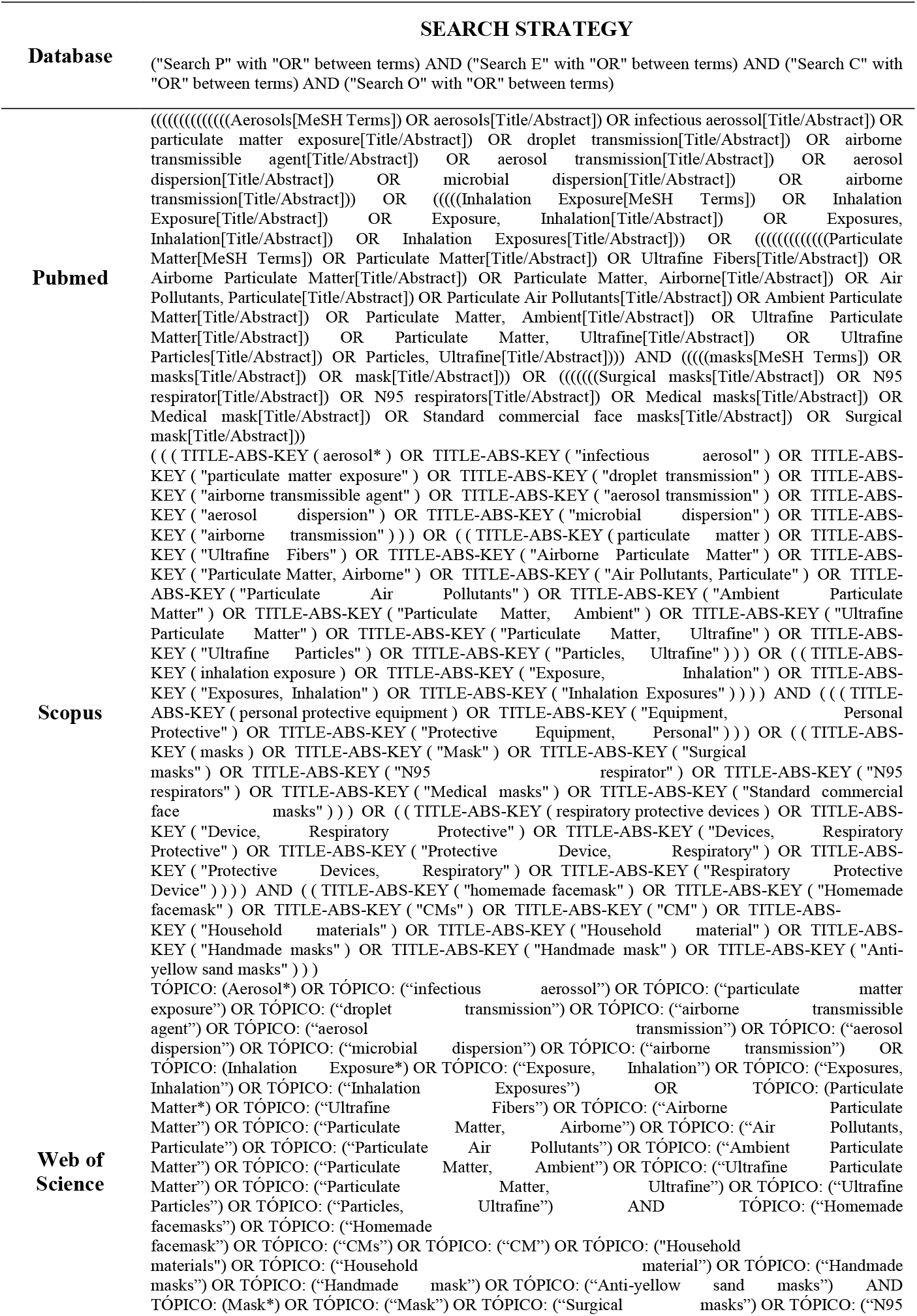
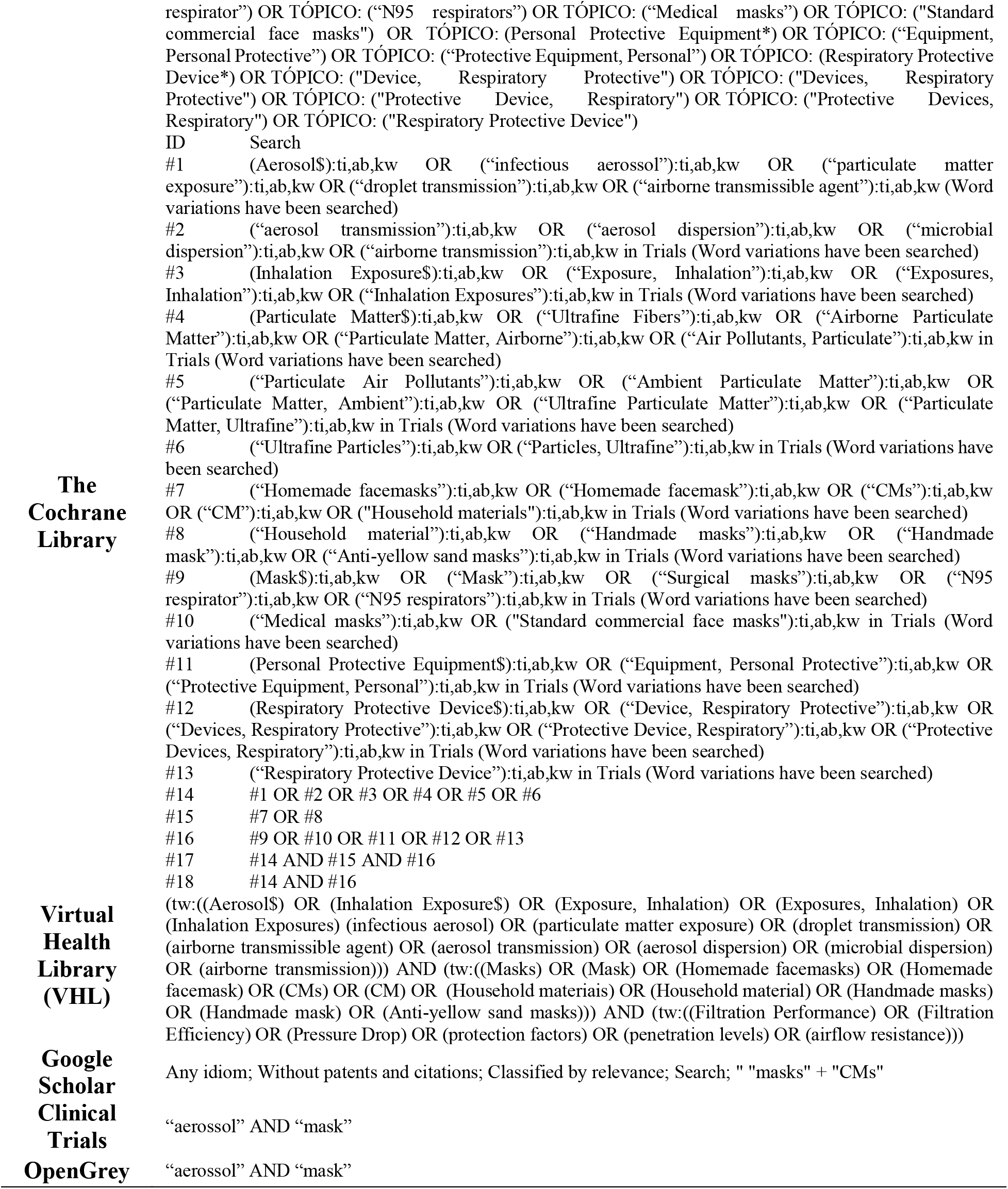
Search Strategy for Each Database.

Inclusion Criteria:

1. Problem: Droplet and/or aerosol dispersion contamination.
2. Exposure: Homemade and/or commercial cloth masks.
3. Comparison: Surgical mask and/or N95 respirator.
4. Outcome: The efficiency of handmade or commercial cloth masks in reducing contamination and the transmission of contaminated droplets and aerosols, by means of laboratory and clinical tests that use surgical masks or N95 respirators for comparison.
5. Study Types: Randomized or non-randomized clinical trials, observational and laboratory studies.

Exclusion Criteria:

1. No comparison group.
2. Case series, opinion articles, animal studies and narrative reviews.

### Data collection

Two authors, independently, sorted the articles by title, abstract and full text, using the bibliographic reference manager Endnote (version X7, Thomson Reuters). Disagreements during study selection and data extraction were settled through a consensus meeting and, when appropriate, by consulting with a third author. The qualitative data extraction table included the following information: Author, Year and Country; Exposure; Comparison; Sample and Method and Authors’ conclusions.

### Data analysis

For all laboratory-based studies evaluated in this systematic review, the Checklist for Quasi-Experimental Studies (non-randomized experimental studies) from The Joanna Briggs Institute (JBI)^16^ was used. The checklist was adapted according to the statements proposed by CRIS Guidelines (Checklist for Reporting *In-Vitro* Studies)^17^, which suggests evaluating factors such as the randomization process, blinding and statistical analysis. The evaluated criteria were divided into seven domains which were categorized with “yes”, “no” or “unclear”. The checklist was individually analyzed for each study and classified as low, moderate or high risk of bias. This final classification was assigned according to the number of domains that presented “no” or “unclear” as an answer. One or two domains were considered as low risk; three or four as moderate risk; and five or more as high risk of bias.

For the evaluation of RoB for the non-randomized clinical trials, the ROBINS-I-tool^18^ was used. The evaluated criteria were divided into pre-intervention, intervention, and post-intervention categories. The RoB was classified as low (one or two domains with “moderate” or “high”), moderate (three or four domains), serious (five or six domains), critical (all the domains), and no information accordingly^19^.

For the randomized clinical trial, the RoB was performed using the Cochrane Collaboration RoB 2.0 tool^20^ which uses the following domains: random sequence generation, allocation concealment, blinding of patients and personnel, blinding of outcome assessor, incomplete outcome data, and selective outcome reporting. Low risk of bias was considered when all key domains were considered at low risk; unclear risk of bias was considered when one or more key domains were unclear and high risk of bias was considered when one or more key domains were considered at high risk.

A meta-analysis was not feasible due to the high methodological heterogeneity identified; however, a detailed qualitative synthesis was performed. and the quality of the evidence of the included studies was performed using GRADE (Grading of Recommendations Assessment, Development and Evaluation – https://gradepro.org/)^21^. The following outcomes were analyzed: Filtration efficiency (%), penetration level (%), airflow resistance, protection factor, cough experiment, pressure drop, surface masks test and occupational health which includes clinical respiratory illness (CRI), influenza-like illness (ILI), laboratory-confirmed respiratory virus infection and pressure differential.

## RESULTS

A total of 2047 records were initially identified in the 8 electronic databases searched: PubMed (n=898), Scopus (n=9), Web of Science (n=7), Cochrane (n=279), Virtual Healthy Library (n=249), OpenGrey (n=2), Google scholar (n=600) and Clinical Trials (n=3). After the removal of 218 duplicates through the Endnote manager, 1829 titles and abstracts were examined. Fifteen records which satisfied the inclusion/exclusion criteria were retained for full text assessment. From those 15 studies, six were excluded: one did not define the type of masks compared^22^; one did not report how the particle penetration rate of the masks compared was obtained^23^; two records examined only factors influencing compliance with the use of medical and cloth masks amongst hospital workers^24,25^; and two evaluated the effectiveness of cloth masks after washing without any comparisons^26,27^ (Table 2).

**Table 2.** List of Excluded Studies (With Reason) After Full Text Review.

| Reference | Reason for exclusion |
| --- | --- |
| Bailey et al., 1968 | No specification of the compared masks |
| Chughtai et al., 2015 | No specification about particle penetration |
| Chughtai et al., 2013 | Study not related with the objective of this Systematic Review |
| Chughtai et al., 2016 | Study not related with the objective of this Systematic Review |
| Kim, 2017 | No comparison with surgical masks or N95 respirator |
| Neupane et al., 2019 | No comparison with surgical masks or N95 respirator |

One additional article was identified after hand search and another was found through a search alert. Finally, 11 articles were selected and included in the qualitative synthesis of this systematic review^11,12,19,28–35^ (Figure 1). The summaries of qualitative and quantitative data are shown in Table 3 and Table 4 respectively. Attempts to communicate by email with corresponding authors were made when data were unavailable. However, only one author responded^19^.

**Table 3.**
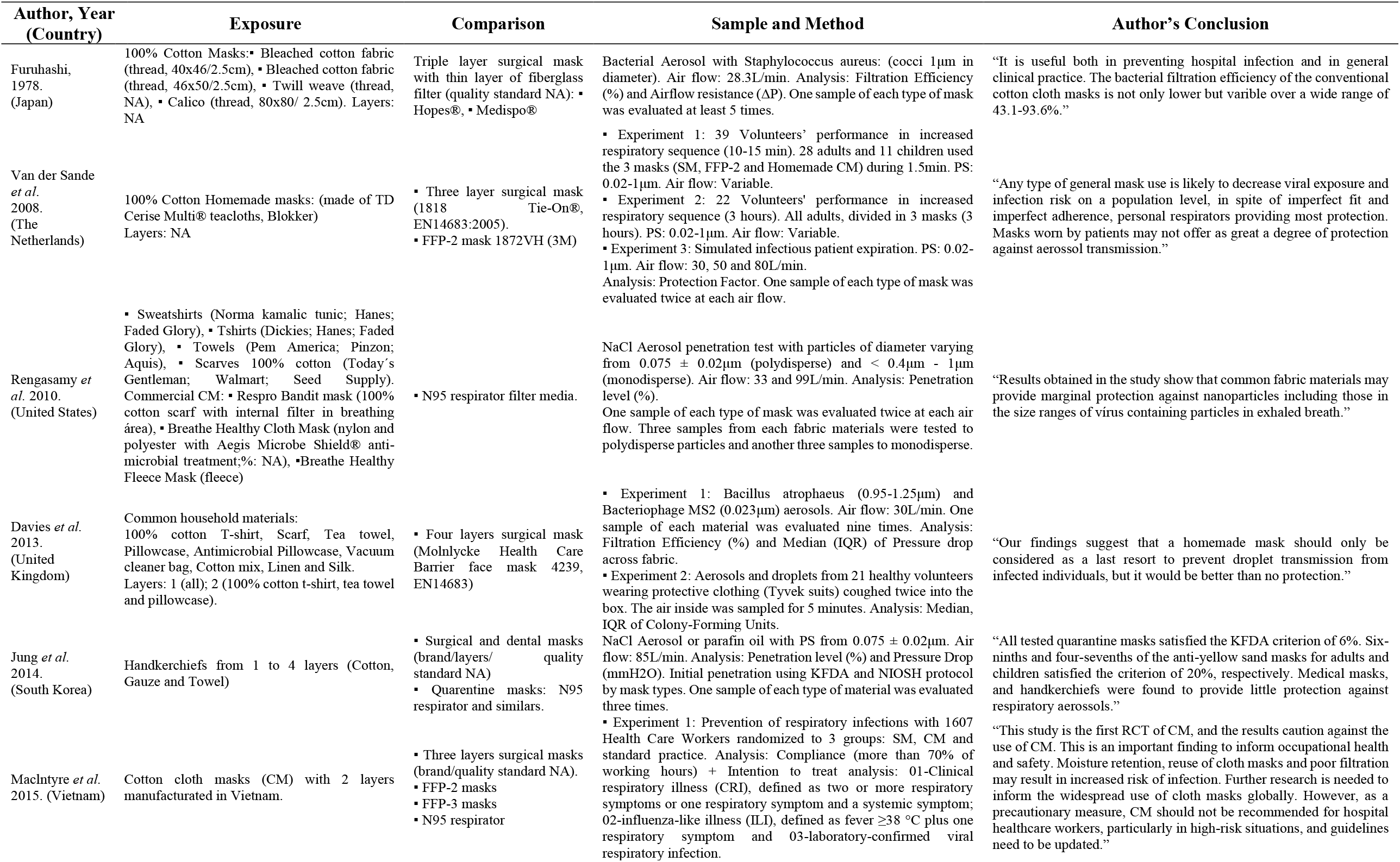
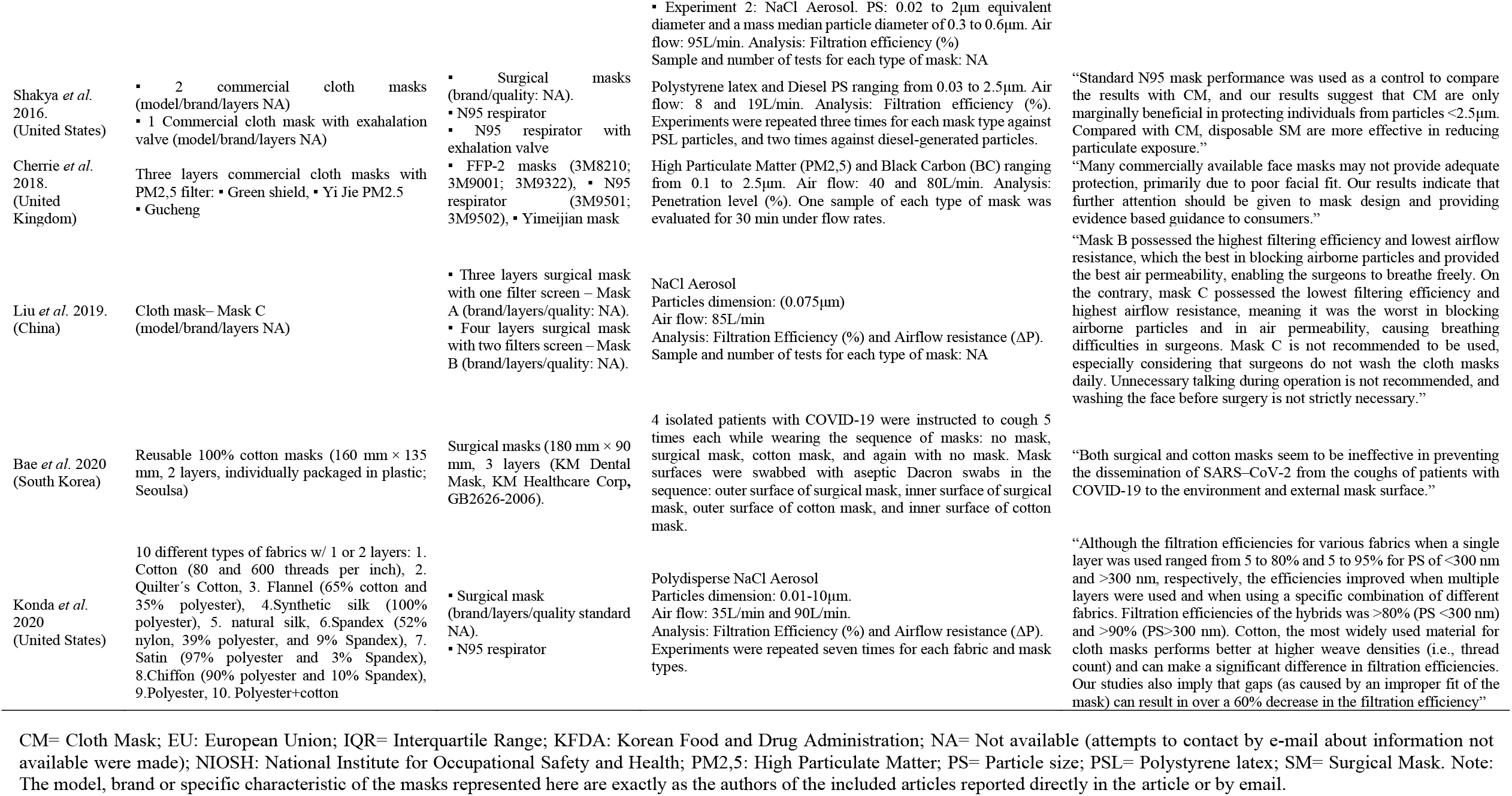
Data Summary From the Studies Included in This Systematic Review.

**Table 4.** Recommended Cloth Masks based on 95% Confidence Interval >60% for Filtration Efficiency and <10% for Penetration Level (Particles Size <0.03 μm).

| Mask (reference) | Particles Size (PS) $\mu\text{m}$ | Air Flow L/min | n | Mean | SD | Lower bound | Upper bound | Recommendation for COVID-19 |
| --- | --- | --- | --- | --- | --- | --- | --- | --- |
| **FILTRATION EFFICIENCY** |  |  |  |  |  |  |  |  |
| CM, Hybrid 1, cotton+chiffon (41) | <0.03 | 35 and 90 | 7 | 97.0 | 2.0 | 95.2 | 98.8 | >90% |
| CM Cotton quilt (41) | <0.03 | 35 and 90 | 7 | 96.0 | 2.0 | 94.2 | 97.8 | >90% |
| CM, Hybrid 2, cotton+silk, no gap (41) | <0.03 | 35 and 90 | 7 | 94.0 | 2.0 | 92.2 | 95.8 | >90% |
| CM, Natural silk, 4 layers (41) | <0.03 | 35 and 90 | 7 | 86.0 | 5.0 | 81.5 | 90.5 | 80-90% |
| CM Vacuum cleaner bag (21) | 0.023 | 30 | 9 | 85.9 | 1.6 | 84.8 | 87.1 | 80-90% |
| CM Hybril 3, cotton+flannel (41) | <0.03 | 35 and 90 | 7 | 85.0 | 2.0 | 83.2 | 86.8 | 80-90% |
| CM Chiffon, 2 layers (41) | <0.03 | 35 and 90 | 7 | 83.0 | 9.0 | 75.0 | 91.0 | 70-90% |
| CM Cotton mix (21) | 0.023 | 30 | 9 | 70.2 | 0.1 | 70.2 | 70.3 | 70-80% |
| CM Cotton, 600TPI, 2 layers (41) | <0.03 | 35 and 90 | 7 | 82.0 | 19.0 | 65.0 | 99.0 | 60-80% |
| CM Antimicrobial pillowcase (21) | 0.023 | 30 | 9 | 68.9 | 7.4 | 63.4 | 74.4 | 60-80% |
| CM inespecified (39) | 0.075 | 85 | NA | NA | NA | NA | NA | Uncertainty |
| CM Linen (21) | 0.023 | 30 | 9 | 61.7 | 2.4 | 59.9 | 63.5 | Uncertainty >50% |
| CM Cotton, 600TPI, 1 layer (41) | <0.03 | 35 and 90 | 7 | 79.0 | 23.0 | 58.4 | 99.6 | Uncertainty >50% |
| CM, Natural silk, 2 layers (41) | <0.03 | 35 and 90 | 7 | 65.0 | 10.0 | 56.1 | 73.9 | Uncertainty >50% |
| CM Tea Towel (21) | 0.023 | 30 | 9 | 72.5 | 22.6 | 55.6 | 89.3 | Uncertainty >50% |
| CM Chiffon, 1 layer (41) | <0.03 | 35 and 90 | 7 | 67.0 | 16.0 | 52.7 | 81.3 | Uncertainty >50% |
| CM Flannel (41) | <0.03 | 35 and 90 | 7 | 57.0 | 8.0 | 49.8 | 64.2 | Not Recommended |
| CM Pillowcase (21) | 0.023 | 30 | 9 | 57.1 | 10.6 | 49.3 | 65.0 | Not Recommended |
| CM, Natural silk, 1 layer (41) | <0.03 | 35 and 90 | 7 | 54.0 | 8.0 | 46.8 | 61.2 | Not Recommended |
| CM 100% Cotton Tshirt (21) | 0.023 | 30 | 9 | 50.9 | 16.8 | 38.3 | 63.4 | Not Recommended |
| CM Scarf (21) | 0.023 | 30 | 9 | 48.9 | 19.8 | 34.1 | 63.6 | Not Recommended |
| CM Silk (21) | 0.023 | 30 | 9 | 54.3 | 29.5 | 32.3 | 76.3 | Not Recommended |
| CM Hybrid 2, cotton+silk, w/ gap (41) | <0.03 | 35 and 90 | 7 | 37.0 | 7.0 | 30.7 | 43.3 | Not Recommended |
| CM Quilter's cotton, 80TPI, 2 layers (41) | <0.03 | 35 and 90 | 7 | 38.0 | 11.0 | 28.2 | 47.8 | Not Recommended |
| **PENETRATION LEVEL** |  |  |  |  |  |  |  |  |
| CM Green shield 3 layers (38) | 0.023 | 30 | 9 | 1-10% | NA | <10% | <10% | <10% |
| CM Yi Jie PM2.5 3 layers (38) | 0.023 | 30 | 9 | 1-10% | NA | <10% | <10% | <10% |
| CM Handkerchief (Gauze 1 layer)- KFDA (19) | 0.075 | 30 | 3 | 99.6 | 0.4 | 98.8 | 100.3 | Not Recommended |
| CM Handkerchief (Gauze 1 layer)- NIOSH (19) | 0.075 | 30 | 3 | 99.3 | 0.3 | 98.7 | 99.9 | Not Recommended |
| CM Handkerchief (Gauze 2 layers)- NIOSH (19) | 0.075 | 30 | 3 | 98.6 | 0.5 | 97.7 | 99.5 | Not Recommended |
| CM Handkerchief (Cotton 1 layer)- NIOSH (19) | 0.075 | 30 | 3 | 98.9 | 0.7 | 97.7 | 100.2 | Not Recommended |
| CM Handkerchief (Cotton 1 layer)- KFDA (19) | 0.075 | 30 | 3 | 98.0 | 0.4 | 97.4 | 98.6 | Not Recommended |
| CM Handkerchief (Gauze 3 layers)- KFDA (19) | 0.075 | 30 | 3 | 98.2 | 0.5 | 97.3 | 99.1 | Not Recommended |
| CM Handkerchief (Gauze 3 layers)- NIOSH (19) | 0.075 | 30 | 3 | 98.0 | 0.4 | 97.3 | 98.7 | Not Recommended |
| CM Handkerchief (Gauze 2 layers)- KFDA (19) | 0.075 | 30 | 3 | 99.0 | 1.0 | 97.2 | 100.9 | Not Recommended |
| CM Handkerchief (Cotton 2 layers)- NIOSH (19) | 0.075 | 30 | 3 | 98.0 | 0.7 | 96.7 | 99.3 | Not Recommended |
| CM Handkerchief (Gauze 4 layers)- KFDA (19) | 0.075 | 30 | 3 | 97.2 | 0.3 | 96.7 | 97.7 | Not Recommended |
| CM Handkerchief (Cotton 3 layers)- NIOSH (19) | 0.075 | 30 | 3 | 96.9 | 0.4 | 96.2 | 97.6 | Not Recommended |
| CM Handkerchief (Gauze 4 layers)- NIOSH (19) | 0.075 | 30 | 3 | 96.4 | 0.4 | 95.7 | 97.0 | Not Recommended |
| CM Handkerchief (Cotton 4 layers)- NIOSH (19) | 0.075 | 30 | 3 | 96.2 | 0.3 | 95.6 | 96.8 | Not Recommended |
| CM Handkerchief (Cotton 2 layers)- KFDA (19) | 0.075 | 30 | 3 | 95.3 | 0.7 | 94.0 | 96.5 | Not Recommended |
| CM Handkerchief (Cotton 3 layers)- KFDA (19) | 0.075 | 30 | 3 | 91.2 | 1.0 | 89.4 | 93.1 | Not Recommended |
| CM Handkerchief (Cotton 4 layers)- KFDA (19) | 0.075 | 30 | 3 | 87.1 | 0.7 | 85.7 | 88.4 | Not Recommended |
| CM Gucheng 3 layers (38) | 0.023 | 30 | 9 | >10% | NA | >10% | >10% | Not Recommended |

**Figure 1.**
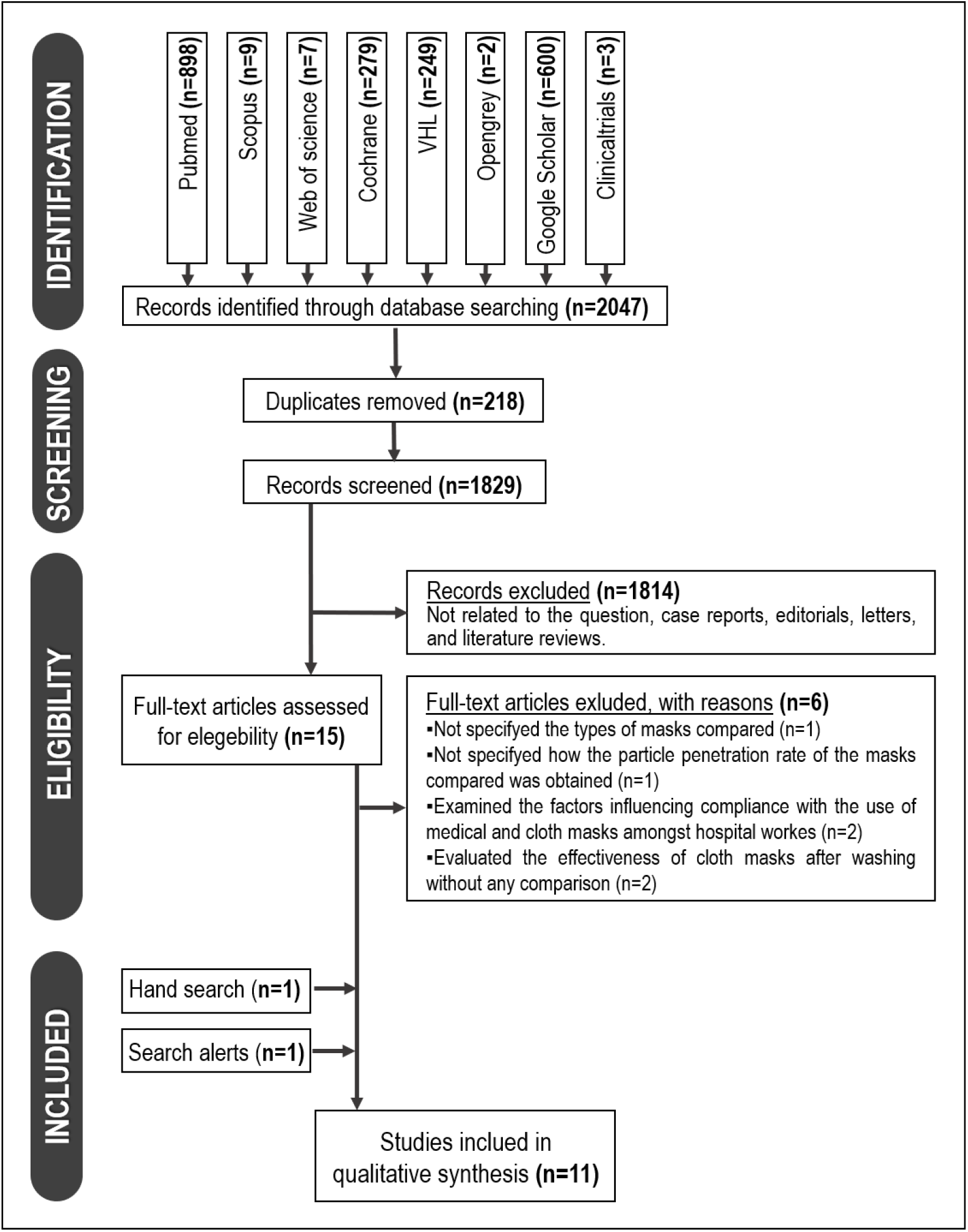
Flowchart with number of records at each stage according to PRISMA statement

According to each type of study, a different tool for assessing RoB was used. From the 11 selected studies for qualitative analysis, there were nine laboratory studies, one non-randomized clinical trial and one randomized clinical trial complemented by laboratory data. RoB was performed separately for each outcome within each study.

An adapted JBI checklist for Quasi-Experimental Studies (experimental studies without random allocation) was applied to ten studies^11,12,19,28–33,35^. Seven domains were evaluated: randomization processes; clearly described methods, interventions, outcome measures; blinding of the assessments; reliable measurement of outcomes and proper statistical analysis (Table 5).

**Table 5.** Risk of Bias of Included Studies According Adapted Quasi Experimental Tool From The Joanna Briggs Institute.

| AUTHOR | DOMAINS |  |  |  |  |  |  | Overall Risk of Bias Judgment |
| --- | --- | --- | --- | --- | --- | --- | --- | --- |
|  | Any randomization process occurs in the study? | Were the methods of the study clearly described? (masks material, particles size and air flow speed) | The masks included in any comparisons received the same intervention? | Were outcome measures of interest taken multiple times? | Were the people assessing the outcomes blinded? | Were outcomes measured in a reliable way? | Was appropriate statistical analysis used? |  |
| Furuhashi (1978) | NO | NO | YES | NO | UNCLEAR | YES | UNCLEAR | HIGH |
| Van der Sande *et al.* (2008) | NO | YES | YES | YES | UNCLEAR | YES | YES | LOW |
| Rengasamy *et al.* (2010) | YES | YES | YES | UNCLEAR | UNCLEAR | YES | UNCLEAR | MODERATE |
| Davies *et al.* (2013) | NO | YES | YES | YES | UNCLEAR | YES | YES | LOW |
| Jung *et al.* (2014) | NO | YES | UNCLEAR | YES | UNCLEAR | YES | YES | MODERATE |
| Macintyre *et al.* (2015), experiment 2. | YES | YES | YES | UNCLEAR | YES | UNCLEAR | YES | LOW |
| Shakya *et al.* (2016) | NO | NO | YES | YES | UNCLEAR | YES | UNCLEAR | MODERATE |
| Cherrie *et al.* (2017) | YES | NO | YES | YES | UNCLEAR | YES | YES | LOW |
| Liu *et al.* (2019). | NO | NO | YES | NO | UNCLEAR | YES | YES | MODERATE |
| Konda *et al.* (2020) | NO | YES | YES | YES | UNCLEAR | YES | YES | LOW |

Only three studies^19,29,30^ reported on the randomization process and one^19^ informed the blinding process, but this investigation was classified as lacking clear information about reliable measurement of outcomes.

Five studies^12,19,30,32,35^ were classified with a low RoB, but only one^30^ reported correctly all the domains, excluding the blinding of the assessments that was unclear. Four studies^11,29,31,33^ were classified with a moderate RoB mainly for not reporting any randomization process and for not clearly describing other domains. Only one study presented a high RoB^28^ because it reported only on the reliable measurement of outcomes and on the interventions.

The ROBINS-I-Tool (Risk of Bias in Non-randomized Studies-of Interventions) was used in one study^34^ that was classified with a high risk of bias (Table 6). The major reason for this RoB rating was due to bias in selection of participants, who had been invited to participate in the research; and bias in classifying interventions since it did not report if the cough velocity was measured and if the patients were under treatment, which can be confounders since the cough velocity and the use of medications can modify the results. In addition, no inclusion and exclusion criteria of participants had been established and this can lead to a heterogeneous sample and unrealistic results. For the cluster randomized trial^19^, RoB was evaluated according to the Cochrane collaboration RoB 2.0 tool, and was rated as low in all domains: random sequence generation, allocation concealment, blinding of patients and personnel, blinding of outcome assessor, incomplete outcome data, and selective outcome reporting (Table 7).

**Table 6.** Risk of Bias of the Included Studies, According to the ROBINS-I Tool.

| AUTHOR | DOMAINS |  |  |  |  |  |  | Overall Risk of Bias Judgment |
| --- | --- | --- | --- | --- | --- | --- | --- | --- |
|  | Bias due to confounding | Bias in selection of participants for the study | Bias in classifying interventions | Bias due to deviations from intended interventions | Bias due to missing data | Bias to measuring outcomes | Bias in selecting reported results |  |
| Bae *et al.* 2020. | Moderate | High | High | Low | Moderate | Moderate | Moderate | High |

**Table 7.**
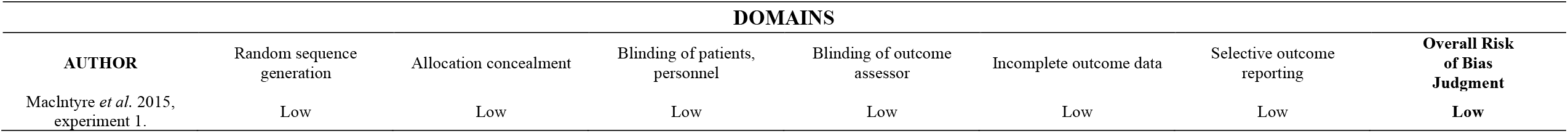
Risk of Bias of the Included Studies, According to Cochrane RoB 2.0.

Regarding the material used in the face masks, six studies evaluated several household materials that could possibly be used for making cloth masks^12,28,29,32,33,35^, while four articles evaluated only commercially available cloth masks^11,19,30,31^. Four studies compared cloth masks with surgical masks only^12,28,31,34^, one compared them with N95^29^ respirators only, one with N95 and FFP-2 respirators^30^, four studies compared cloth masks with both surgical masks and N95 respirators or similar (FFP-2)^11,32,33,35^, and only one study compared cloth masks with surgical masks, and FFP-2 and FFP-3 respirators^19^ (Table 3).

Regarding the experimental model, performed by simulation, five studies used NaCl aerosol with particles size reported: 0.075μm^29,31,33^, 1μm^29^, 0.02 to 2μm^19^ and 10nm to 10 μm^35^. The air flow speed in these studies was 33L/min and 99L/min^29^, 85L/min^31,33^, 95L/min^19^ and in the last study two velocities 35 L/min and 90 L/min^41^ were used. Two studies used microbial aerosols: one was contaminated aerosol with *Staphylococcus aureus* of 1μm in diameter at 28L/min air flow speed^28^, one used *Bacillus atrophaeus* with 0.95-1.25μm and *Bacteriophage MS2* with 0.023μm, both with a 30L/min air flow speed^12^. One study^11^ used Polystyrene latex and Diesel Particles from 0.03 to 2.5μm in an air flow speed of 8 and 19L/min; and one study^30^ used high particulate matter from 0.1 to 2.5μm with 40 and 80L/min. Three studies evaluated more than one outcome, and also used volunteers^12,19,32^. One study^34^ did not perform an experiment by simulation and concluded that both surgical and cloth masks are inefficient in containing the spread.

### Anti-contamination measurements

For anti-contamination measurements, the filtration efficiency (%) was evaluated by six studies^11,12,19,28,31,35^, where three of them compared cloth masks with surgical masks only^12,28,31^. The first one^12^ analyzed several homemade cloth masks and found that better results were achieved by the tea towel with 2 layers (96.71 ± 8.73) and vacuum cleaner bag (94.35 ± 35), with results similar to surgical masks (96.35 ± 0.68). Cotton mix (74.60 ± 11.17) and 100% cotton T-shirts with 2 layers (70.66 ± 6.83) also presented good results, while linen (60.00±11.18) and silk (58.00 ± 2.75) had the worst results. The second one^12,28^ analyzed three different cloth masks and reported that the best result was achieved by the twill cloth mask(93.6±1.16) with no difference compared to Hopes® surgical mask (98.1±1.02, p<0.05), and the last one^31^ concluded that the surgical mask with two filter screens presented 60-80% of filtration efficiency while cloth masks about 20%.

Three studies^11,19,35^ compared cloth masks with both surgical masks and N95 respirators or similar. The first one^11^ reported that the efficiency of cloth masks presented the worst results (39% to 65%) in comparison to the other two groups, the second study^19^ noted penetration of particles through the cloth masks to be very high (97%), but neither study reported the fabric of the cloth masks. The last one^35^ found that hybrid fabrics potentially provide protection against the transmission of aerosol particles, with a filtration efficiency of three types of hybrid fabrics: cotton/chiffon (97 ± 1), cotton/silk (94 ± 2) with no gap, and cotton/flannel (95 ± 2) even better than N95 respirators (85±15) in relation to <300 nm particles.

The penetration level (%) was measured by three studies^29,30,33^. The first one^33^ compared cloth masks with surgical masks and N95 respirators and found a high penetration level in handkerchiefs mainly made of gauze and with one layer (99.57 ± 0.40), and better results were found in a certified N95 respirator group with a penetration level (0.62 ± 0.36). The remaining two^29,30^ compared the cloth masks only with N95 and/or FFP-2 respirators and both of them noted a high penetration level in cloth masks in relation to comparison group. One^29^ found better results between the cloth fabrics in sweatshirts with 40% of penetration level at 33L/min and 57% at 99L/min. The other^30^ found that a mask named Yi Jie PM2.5 presented the lowest degree of penetration between the other cloth masks options with 67.3% (IQR: 56.6%,75.2%), but even so with worse results when compared to the best N95 brand 3M9322 1.8% (IQR: 0.6%,4.7%).

Recommended Cloth Masks based on 95% Confidence Interval >60% for Filtration Efficiency and <10% for Penetration Level (Particles Size <0.03 μm) are reported in Table 4.

Occupational health was evaluated by only one study^19^ and the rates of clinical respiratory illness (CRI), influenza-like illness (ILI) and laboratory-confirmed virus infections were higher in the cloth mask arm compared to medical masks, mainly ILI, with a relative risk =13.00 (95% CI 1.69,100.07).

Protection factor^32^ showed that surgical masks provided about twice as much protection as homemade masks, FFP2 masks provided 50 times as much protection as homemade masks, and 25 times as much protection as surgical masks.

### Anti-transmission measurements

The protection factor of cloth, surgical and FFP-2 masks were evaluated by one study^32^ which showed that cloth masks presented a considerably lower protection factor (1.9, CI95% 1.5,2.3) especially in children. Protection offered by a surgical mask and FFP2 respirator did not differ.

Two studies^12,34^ evaluated particle dissemination when coughing. The first one^12^ found that both surgical and cloth masks reduced the total number of microorganisms expelled when coughing in comparison with coughing without a mask, while the second one^34^ found that neither cloth or surgical masks effectively filtered the virus expelled when coughing.

### Breathability

Studies evaluating pressure drop (PD)^12,33^ and airflow resistance (Pa)^28,31^ had demonstrated that tea towel^12^, vacuum cleaner bag masks^12^, cotton handkerchief with four layers^33^, twill weave^28^ and bleached cotton^28^ had greater potential to block contaminated patient particles outside the cloth mask. However, they can cause a suffocating sensation to the user.

On the other hand, some of the evaluated fabrics presented a good breathability, such as calico^28^, silk^12^, linen^12^, cotton and gauze handkerchief^33^. One study^35^ reported that the average differential pressure across all of the fabrics studied at a flow rate of 1.2 CFM was found to be 2.5 (0.4) Pa, indicating conditions for good breathability, but we can’t claim that these cloth masks are able to contain or reduce particles expelled by the user.

### Quality of the evidence

GRADE assessment was divided into anti-contamination and anti-transmission and breathability outcomes. For the outcomes included in the anti-contamination the quality of the evidence ranged from low to moderate level (Occupational health). For the anti-transmission and breathability outcomes, the quality of the evidence ranged from very low to moderate due to the bias of the included studies and magnitude of effect (Tables 8 and 9).

**Table 8.** GRADE of Anti-Contamination Measurements.

| Outcomes | Impact | Nº of participants (studies) | Certainty of the evidence (GRADE) |
| --- | --- | --- | --- |
| Filtration Efficiency (%) - Cloth Masks X Surgical Masks | Three studies evaluated the comparison between cloth masks and surgical masks. All concluded that surgical masks have greater filtration efficiency than cloth masks. One presented high risk of bias, one presented moderate and one low risk. | 3 laboratory setting studies<sup>21,36,39</sup> | ⊕⊕○○<br>LOW *†‡ |
| Filtration Efficiency (%) - Cloth Masks X Surgical Masks X Respirator (N95 and/or FFP-2) | Three studies evaluated the comparison between cloth masks and respirators (N95 and/or FFP-2). One study reported that N95 and/or FFP-2 respirators have greater filtration efficiency, followed by the surgical and finally cloth masks, one found that N95 respirators presented better results, but one of the cloth masks (with a valve) presented similar filtration efficiency to surgical masks and one found similar results between the three groups. Two presented moderate risk and one low risk. | 3 laboratory setting studies<sup>20,22,41</sup> | ⊕⊕○○<br>LOW *†‡ |
| Penetration level (%) - Cloth Masks X Respirator (N95 and/or FFP-2) | Two studies evaluated the comparison between cloth masks, N95 and/or FFP-2. One study concluded that penetration levels were much higher in the cloth masks and one study concluded that results were similar in both groups when analyzing two of the three cloth masks. One study presented low risk of bias and the other a moderate risk. | 2 laboratory setting studies<sup>37,38</sup> | ⊕⊕○○<br>LOW †‡ |
| Penetration level (%) - Cloth masks X Surgical masks X Respirator (N95 and/or FFP-2) | One study evaluated the comparison of the penetration level between cloth masks, surgical masks and N95 respirators (or similar). N95/FFP-2 respirator groups presented the better results, followed by the surgical masks and the cloth masks. The study presented moderate risk of bias. | 1 laboratory setting study<sup>19</sup> | ⊕⊕○○<br>LOW *‡ |
| Protection Factor - Cloth Masks X Surgical Masks X FFP-2 | One study with a low risk of bias evaluated the protection factor in anti-contamination between cloth masks, surgical masks and FFP-2 respirators and concluded that the surgical and FFP-2 masks presented a higher protection factor than the cloth masks. | 1 laboratory setting study<sup>13</sup> | ⊕⊕⊕○<br>MODERATE *‡ |
| Occupational Health - Cloth Masks X Surgical Masks X Control | One study with low risk of bias evaluated the occupational health of health workers and found that rates of clinical respiratory illness, influenza-like illness and laboratory-confirmed virus infections were lowest in the medical mask arm, followed by the control arm, and highest in the cloth mask arm. | 1 randomized clinical trial<sup>22</sup> | ⊕⊕⊕○<br>MODERATE † |

\*Furuhashi et al., Van der Sande et al., Davies et al., Jung et al., Shakya et al., Liu et al., Konda et al., didn't present any randomization process in the study; †Furuhashi et al., Mecintyre et al., Shakya et al., Cherrie et al., and Liu et al., failed to describe the study methods; ‡Furuhashi et al., Van der Sande et al., Rengasamy et al., Davies et al., Jung et al., Shakya et al., Cherrie et al., and Liu et al., probably didn't blind the people assessing the outcome.

**Table 9.**
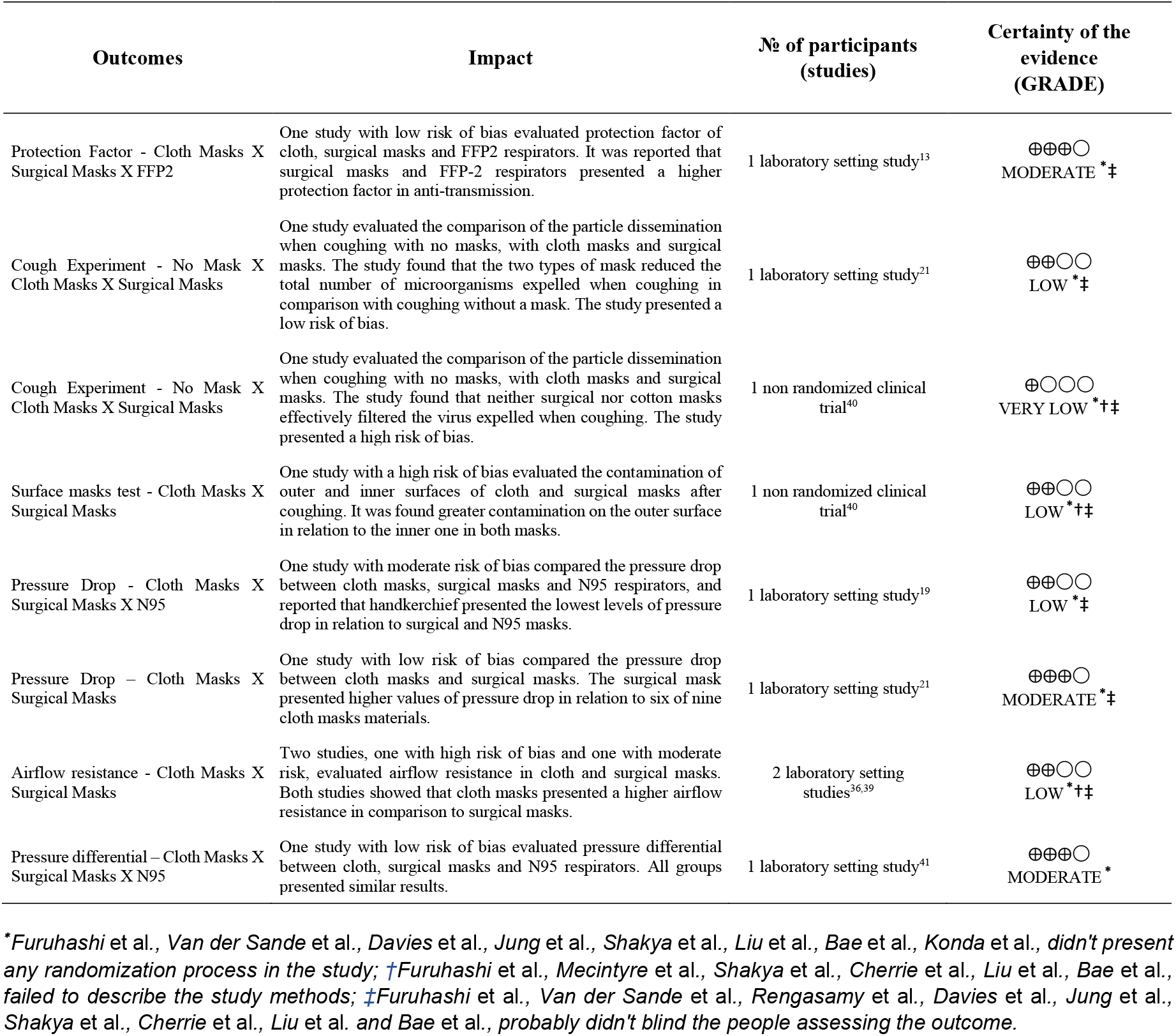
GRADE of Anti-Transmission and Breathability Measurements.

In general terms, surgical and N95 and/or FFP-2 masks presented better results in most of the factors evaluated in comparison with the cloth masks, with very low to moderate certainty of evidence level depending on the outcome analyzed. Regardless of some benefits for cloth mask users, the results are hard to summarize and generalize because of the variety of fabrics and layers evaluated.

These results should be viewed with caution given the quality of the evidence and the fact that almost all the included studies evaluated the outcome of interest in a laboratory setting. Furthermore, elements of statistical precision between the groups are scarce, and outcomes such as degree of protection, pressure drop, surface masks test and occupational health were each evaluated in only one study.

## DISCUSSION

Our results suggest that cloth masks present worse outcomes for filtration efficiency, penetration level and protection factor in comparison with medical masks, when evaluated in a laboratory-based examining small particles. In accordance with other study^8^, these results seem to substantially underestimate the efficiency of cloth masks for source control in real life when referring to blocking droplets ejected by the wearer, since in most cases the particles used in these studies were smaller than a droplet and generally ranged between 5 μm to 10 μm^36,37^. For this reason, it is suggested that the use of cloth masks by the general public is likely a useful public health measure in reducing COVID-19 contamination and transmission. In addition, the fact that cloth masks are not as effective as surgical masks does not mean that they provide no protection. Anything that contributes to controlling the spread of a virus should be encouraged from a population-based point of view. Multiple approaches that alone do not a have a major impact when combined could have a multiplicative effect in slowing the spread of a virus like COVID-19 by reducing the transmission rate.

Overall, the filtration efficiency of the fabric depends on a variety of factors: the composition of the fabric and some characteristics of the particles to which it is exposed such as their size and velocity. These factors are fundamental to evaluate the quality of the masks. Only seven studies^11,12,29–33^ presented particle sizes compatible with the new coronaviruses (0.06-0.14 μm)^38^. This lack of complete information directly affected the potential bias of these studies. If the particle sizes were known, we could have better evaluated the efficiency of cloth masks for the general population against the coronavirus.

Only one study^34^ evaluated the use of cloth masks in patients with COVID-19, but it is important to clarify that this study presented some limitations such as a small sample size and inconsistent data, including no detection of viral load in one participant’s cough test (including without a mask) and no detection of viral load in the inner surface of the masks in three of four patients after coughing. The other studies assessed other types of bacteria and viruses, but this did not seem to affect the results. The study that evaluated the use of cloth masks by healthcare workers^19^ did not recommend their use by these professionals. A recent systematic review^39^ showed that low quality evidence was presented in studies evaluating the use of PPE, face masks (surgical and N95) and eye protection to prevent infectious diseases in healthcare workers. The authors highlighted the urgent need for randomized clinical trials with better methodological quality. However, results in a healthcare setting are not readily generalizable to the population where any measure, even not as efficient as a measure in a healthcare setting, can provide some source control.

Another recent systematic review^14^ investigated physical distancing, face masks and eye protection to prevent person-to-person transmission of COVID-19 and they supported that physical distancing of at least 1 m is strongly associated with protection, but distances up to 2 mm might be more effective. Regarding the use of face masks, it was found that it could result in a large reduction in risk of infection, with stronger association with N95 or similar respirators when compared with surgical masks. Eye protection also was associated with less infection.

Among the studies that reported the materials used to fabricate cloth masks, the vacuum cleaner bag presented good results, but it’s important to clarify that this material has a high pressure drop, rendering it unsuitable for a face mask, therefore the use of tea towel was recommended instead^12^. In relation to layers of masks, the number used seems to be directly proportional to the filtration capacity in most of the laboratory studies and could be a solution to improve the results achieved by cloth masks^40–43^. The combination of various commonly available fabrics can potentially provide significant protection against the contamination of aerosol particles, as a hybrid of cotton and silk mask seems to present results of filtration efficiency very similar to surgical and N95 masks^35^.

Cloth masks can be effective depending on the fabric and number of layers used. It could decrease the air passage from inside to outside of the masks, thereby favoring the decrease of the microorganisms expelled during speaking, coughing, or sneezing. However, it is critical that it is well adapted to the facial contour, since the presence of gaps caused by an improper fit of the mask can result in over a 60% decrease of their filtration efficiency^35^. In addition, some authors recommended that in situations of public emergency, with limited evidence, mechanistic and analogous evidence and professional judgment become important. In these cases, the use of facial masks, along with other health measures, such as personal hygiene, can help mitigate the COVID-19 epidemic^44^.

A recent rapid systematic review^45^ evaluated the use of medically manufactured facemasks and similar barriers to prevent respiratory illness such as COVID-19. According to the RCTs, the results showed that the use of facial masks may present little protection against primary infections through casual contact with the community, and modestly protect against domestic infections when infected and non-infected members wear face masks. In observational studies the evidence in favors of wearing facemasks was stronger. This is an important point to be cited since the clinical studies could often suffer from poor compliance and controls using facemasks^45^. Therefore, the correct and continuous use of these protections by the public could improve the clinical results.

This fact can also be supported by a mathematical modelling study that described the spread of COVID-10 infection. Modelling studies suggest that if most people wear masks, the transmission rate can decrease to 1.0^9^. Moreover, cloth masks could be an additional tool to enhance awareness of the importance of physical distancing in public places, serving as a visual reminder^8^.

Another study^46^ discussed the potential effectiveness of the universal adoption of homemade cloth facemasks. They found that that the growth rate of deaths in countries without mask norms is 21%, while in countries with such norms is 11%. Although researchers may disagree on the magnitude of the reduction in transmissibility, the benefits found can be highly expressive and beneficial to the transmission and control process of the disease^8^. Public use of facemask may increase awareness regarding the disease among the population and can contribute to the reduction of the transmission rates.

It is well-known that the virus may survive on the surface of face masks^47^, and contamination may occur since the cloth mask may transfer pathogen to bare hands during the repeated donning and doffing^17^, so it’s very important to wash hands as much as possible and wash the masks daily. Conversely, a study^21^ showed that washing and drying practices could drop by 20% the filtering efficiency of cloth masks after the 4th cycle, due to the increase of the pore size and the expansion of the fabric. It is important to highlight that the masks were air dried to make sure that the cloth fibers were not stretched out, since stretching cloth masks surface also altered the pore size and potentially decreased the filtering efficiency. Further studies about wash and dry care of cloth masks are needed to obtain a longer durability with efficiency. Moreover, authorities need to provide clear guidelines for the use, cleaning, and reuse of facemasks.

A guideline from the WHO^48^ encourage the use of respiratory hygiene in all people with acute respiratory infections (ARIs) and it includes the use of medical or cloth masks. Although the quality of evidence has been considered very low^2^, there was consensus that the advantages of the use of respiratory hygiene and an assessment of values and preferences provided sufficient basis for the strong recommendation. Thus, the importance of cloth masks use by the general population seems an effective way of source control, as people in a pre-symptomatic phase can already spread the virus. This is a simple and low-cost measure that in conjunction with other strategies can be extremely helpful in control and mitigation of the disease.

This systematic review identified some limitations in the primary studies and only two of the included studies were clinical trials. A more realistic comparison between groups was hampered by the lack of detailed features information on the masks studied. There was a lack of studies comparatively assessing the various fabrics, utilizing different particles sizes and designs of cloth masks, and taking into consideration the importance of a good fit on effectiveness. Besides that, new studies with a better methodological quality and randomized clinical trial specifically related to COVID-19 are in urgent needed.

## CONCLUSION

Cloth masks seem to provide some degree of protection against contamination and transmission by droplets and aerosols. It is suggested that the use of cloth masks by the public is a useful public health measure that can protect the wearer and at the same time act as source decrease disease transmission. However, according to one RCT, cloth masks should not be recommended for healthcare workers. Based on very low to moderate quality of evidence the level of efficiency of cloth masks is difficult to generalize because of the variety of fabrics and layers evaluated, the efficiency is higher when cloth masks are made of hybrid fabrics with multiple layers.

## Data Availability

Not applicable.

## Acknowledgments

We thank Dr. Sandra Meilhubers for comments and pre-submission review. There was no funding source for this study.

## REFERENCES

1. World Health Organization [homepage] Preventing epidemics and pandemics. Available from: https://www.who.int/activities/preventing-epidemics-and-pandemics (accessed April 2, 2020).

2. Jefferson T, Del Mar C, Dooley L, Ferroni E, Al-Ansary LA, Bawazeer GA et al. Physical interventions to interrupt or reduce the spread of respiratory viruses. Cochrane database Syst Rev 2011; 7: CD006207.

3. American Enterprise Institute. National coronavirus response: A road map to reopening. [homepage] March 24, 2020. Available from: https://www.aei.org/research-products/report/national-coronavirus-response-a-road-map-to-reopening/ (accessed April 2, 2020).

4. He X, Lau EH, Wu P, Deng X, Wang J, Hao X, et al. Temporal dynamics in viral shedding and transmissibility of COVID-19. Nat Med 2020; 26: 672–5.

5. Nishiura H, Linton N, Akhmetzhanov A. Serial interval of novel coronavirus (COVID-19) infections. IJID 2020; 284–286.

6. Godoy LRG, Jones AE, Anderson TN, Fisher CL, Seeley KML, Beeson EA, et al. Facial protection for healthcare workers during pandemics: a scoping review. BMJ Global Health 2020; 5: e002553.

7. Centers for Disease Control and Prevention. [homepage] Coronavirus Disease 2019 (COVID-19) - Prevent Getting Sick - Protect Yourself. April 24, 2020. Available from:https://www.cdc.gov/coronavirus/2019-ncov/prevent-getting-sick/prevention.html (accessed April 2, 2020).

8. Howard J, Huang A, Li Z, Tufekci Z, Zdimal V, van Der Westhuizen HM, et al. Face masks against COVID-19: an evidence review. 2020. doi:10.20944/preprints202004.0203.v1 (preprint)

9. Greenhalgh T, Howard J. Masks for all? The science says yes. [homepage] April 13, 2020. Available from: https://www.fast.ai/2020/04/13/masks-summary/ (accessed April 2, 2020).

10. Liu Y, Funk S, Flasche S. The contribution of pre-symptomatic infection to the transmission dynamics of COVID-2019. Wellcome Open Res 2020; 5: 58.

11. Shakya K, Noyes A, Kallin R, Peltier R. Evaluating the efficacy of cloth facemasks in reducing particulate matter exposure. Journal of exposure science & environmental epidemiology 2017; 27: 352–7.

12. Davies A, Thompson K, Giri K, Kafatos G, Walker J, Bennett A. Testing the efficacy of homemade masks: would they protect in an influenza pandemic? Disaster medicine and public health preparedness 2013; 7: 413–8.

13. Greenhalgh T, Schmid M, Czypionka T, Bassler D, Gruer L. Face masks for the public during the covid-19 crisis. BMJ 2020; 369.

14. Chu DK, Akl EA, Duda S, Solo K, Yaacoub S, Schünemann HJ, et al. Physical distancing, face masks, and eye protection to prevent person-to-person transmission of SARS-CoV-2 and COVID-19: a systematic review and meta-analysis. The Lancet 2020. Published online June 1. DOI: https://doi.org/10.1016/S0140-6736(20)31142-9

15. Moher D, Shamseer L, Clarke M, Ghersi D, Liberati A, Perricrew M, et al. Preferred reporting items for systematic review and meta-analysis protocols (PRISMA-P) 2015 statement. 2015; 4: 1.

16. Tufanaru C, Munn Z, Aromataris E, Campbell J, Hopp L. Chapter 3: Systematic reviews of effectiveness. In: Aromataris E, Munn Z (Editors). Joanna Briggs Institute Reviewer’s Manual: The Joanna Briggs Institute.; 2017.

17. Krithikadatta J, Gopikrishna V, Datta M. CRIS Guidelines (Checklist for Reporting In-vitro Studies): a concept note on the need for standardized guidelines for improving quality and transparency in reporting in-vitro studies in experimental dental research. J Conserv Dent 2014; 17: 301.

18. Sterne J, Hernán M, Reeves B, Savocic J, Berkman ND, Viswanathan M, et al. ROBINS-I : a tool for assessing risk of bias in non-randomised studies of interventions.. BMJ 2016; 355: i4919–i.

19. MacIntyre C, Seale H, Dung T, Hien NT, Nga PT, Chughtai AA, et al. A cluster randomised trial of cloth masks compared to medical masks in healthcare workers. BMJ Open 5: e006577. BMJ Open 2015; 5: e006577.

20. Higgins JP, Sterne JA, Savovic J, Page MJ, Hróbjartsson A, Bourton I, et al. A revised tool for assessing risk of bias in randomized trials.. Cochrane Database of Syst Rev 2016; 10 (Suppl 1): 29–31.

21. Balshem H, Helfand M, Schünemann H, Oxman AD, Kunz R, Brozek J, et al. GRADE guidelines: 3. Rating the quality of evidence. J Clin Epidemiol 2011; 64: 401–6.

22. Bailey R, Giglio P, Blechman H, Nunez C. Effectiveness of disposable face masks in preventing cross contamination during dental procedures. Journal of dental research 1968; 47: 1062–5.

23. Chughtai A, MacIntyre C, Ashraf M, Zheng Y, Yang P, Wang Q, et al. Practices around the use of masks and respirators among hospital health care workers in 3 diverse populations. Am J Infect Control 2015; 43: 1116–8.

24. Chughtai A, Seale H, MacIntyre C. Availability, consistency and evidence-base of policies and guidelines on the use of mask and respirator to protect hospital health care workers: a global analysis. BMC Res Notes 2013; 6: 216.

25. Chughtai AA, Seale H, Dung TC, Hayen A, Rahman B, Raina MacIntyre C. Compliance with the Use of Medical and Cloth Masks Among Healthcare Workers in Vietnam. Ann Occup Hyg 2016; 60: 619–30.

26. Kim SW. Changes of Particle Filtration Efficiency of Cloth Masks by Machine Washing and Cloth Expansion. J Korean Soc Occup Environ Hyg 2017; 27: 115–22.

27. Neupane B, Mainali S, Sharma A, Giri B. Optical microscopic study of surface morphology and filtering efficiency of face masks. PeerJ 2019; 7: e7142.

28. Furuhashi M. A study on the microbial filtration efficiency of surgical face masks--with special reference to the non-woven fabric mask. Bull Tokyo Med Dent Univ 1978; 25: 7–15.

29. Rengasamy S, Eimer B, Shaffer R. Simple respiratory protection - Evaluation of the filtration performance of cloth masks and common fabric materials against 20-1000 nm size particles. Ann Occup Hyg 2010; 54: 789–98.

30. Cherrie J, Apsley A, Cowie H, Steinle S, Mueller W, Lin C, et al. Effectiveness of face masks used to protect Beijing residents against particulate air pollution. Ann Occup Environ Med 2018; 75: 446–52.

31. Liu Z, Yu D, Ge Y, Wang L, Zhang J, Li H, et al. Understanding the factors involved in determining the bioburdens of surgical masks. Ann Transl Med 2019; 7: 754.

32. van der Sande M, Teunis P, Sabel R. Professional and home-made face masks reduce exposure to respiratory infections among the general population. PloS one 2008; 3: e2618.

33. Jung H, Kim J, Lee S, Lee J, Kim J, Tsai P, et al. Comparison of filtration efficiency and pressure drop in anti-yellow sandmasks, quarantine masks, medical masks, general masks, and handkerchiefs. Aerosol Air Qual Res 2014; 14: 991–1002.

34. Bae S, Kim M, Kim J, Cha HH, Lim JS, Jung J, et al. Effectiveness of Surgical and Cotton Masks in Blocking SARS–CoV-2: A Controlled Comparison in 4 Patients. Ann Intern Med 2020. https://doi.org/10.7326/M20-1342.

35. Konda A, Prakash A, Moss G, Schmoldt M, Grant G, Guha S. Aerosol Filtration Efficiency of Common Fabrics Used in Respiratory Cloth Masks. ACS Nano 2020; 14, 6339–6347 https://doi.org/10.1021/acsnano.0c03252.

36. Morawska L, Johnson G, Ristovski Z, Hargreaves M, Mengersen K, Corbett S, et al. Size distribution and sites of origin of droplets expelled from the human respiratory tract during expiratory activities. J Aerosol Sci 2009; 40: 256–69.

37. Duguid J. The size and the duration of air-carriage of respiratory droplets and droplet-nuclei. J Epidemiology Infection 1946; 44: 471–9.

38. Wang C, Liu Z, Chen Z, Huang X, Xu M, He T, et al. The establishment of reference sequence for SARS-CoV-2 and variation analysis. J Med Virol 2020. doi:10.1002/jmv.25762 (preprint).

39. Verbeek JH, Rajamaki B, Ijaz S, Tikka C, Ruotsalainen JH, Edmond MB, et al. Personal protective equipment for preventing highly infectious diseases due to exposure to contaminated body fluids in healthcare staff. Cochrane Database of Syst Rev 2019; 7: CD011621.

40. Kellogg WH, MacMillan G. An experimental study of the efficacy of gauze face masks. Am J Public Health 1920; 10: 34–42.

41. Leete HM. Some experiments on masks. Lancet 1919; 193: 392–3.

42. Haller DA, Colwell RC. The protective qualities of the gauze face mask: experimental studies. JAMAAP 1918; 71: 1213–5.

43. Weaver GH. Droplet infection and its prevention by the face mask. J Infect Dis 1919; 24: 218–30.

44. Chan K, Yuen K. COVID-19 epidemic: disentangling the re-emerging controversy about medical facemasks from an epidemiological perspective. Int J Epidemiol 2020. DOI: 10.1093/ije/dyaa044 (preprint).

45. Brainard J, Jones N, Lake I, Hooper L, Hunter P. Facemasks and similar barriers to prevent respiratory illness such as COVID-19: A rapid systematic review. medRxiv 2020. DOI: https://doi.org/10.1101/2020.04.01.20049528 (preprint).

46. Abaluck J, Chevalier JA, Christakis NA, Forman HP, Kaplan EH, Ko, A et al. The Case for Universal Cloth Mask Adoption and Policies to Increase Supply of Medical Masks for Health Workers. SSRN Scholarly Paper 2020: ID 3567438. available at SSRN: http://dx.doi.org/10.2139/ssrn.3567438

47. Osterholm M, Moore K, Kelley N, Brosseau LM, Wong G, Murphy FA, et al. Transmission of Ebola viruses: what we know and what we do not know. MBio 2015; 6: e00137–15.

48. World Health Organization. Infection prevention and control of epidemic-and pandemic-prone acute respiratory infections in health care: World Health Organization; 2014.

